# Visceral Leishmaniasis-HIV coinfection as a predictor of increased *leishmania* transmission at the village level in Bihar, India

**DOI:** 10.1101/2020.09.24.20200709

**Authors:** Kristien Cloots, Pia Marino, Sakib Burza, Naresh Gill, Marleen Boelaert, Epco Hasker

**Affiliations:** Department of Public Health, Institute of Tropical Medicine, Antwerp, Belgium; Erasmus Mundus Joint Master Degree Infectious Diseases & One Health Programme; Médecins Sans Frontières, New Delhi, India; National Vector Borne Disease Control Programme, India

## Abstract

**Background:** **V**isceral leishmaniasis (VL) is on the verge of being eliminated as a public health problem in the Indian subcontinent. Although Post-kala-azar dermal leishmaniasis (PKDL) is recognized as an important reservoir of transmission, we hypothesized that patients with VL co-infected with Human Immunodeficiency Virus (HIV) may also be important reservoirs of sustained leishmania transmission. We therefore investigated to what extent cases of PKDL or VL-HIV are associated with VL incidence at the village level in Bihar, India.

**Methods:** VL, VL-HIV, and PKDL case data from six districts within the highly VL-endemic state of Bihar, India were collected through the Kala-Azar Management Information System for the years 2014 – 2019. Multivariate analysis was done using negative binomial regression controlling for year as a fixed effect and block (subdistrict) as a random effect.

**Findings:** Presence of VL-HIV and PKDL cases were both associated with a more than twofold increase in VL incidence at village level, with Incidence Rate Ratios (IRR) of 2.16 (95% CI 1.81 – 2.58) and 2.37 (95% CI 2.01 – 2.81) for VL-HIV and PKDL cases respectively. A sensitivity analysis showed the strength of the association to be similar in each of the six included subdistricts.

**Conclusions:** These findings indicate the importance of VL-HIV patients as infectious reservoirs, and suggest that they represent a threat equivalent to PKDL patients towards the VL elimination initiative on the Indian subcontinent, therefore warranting a similar focus.

**CONTRIBUTION TO THE FIELD:** Visceral leishmaniasis (VL) – also called kala azar on the Indian subcontinent - is a parasitic disease which is fatal if not treated timely. Since the elimination initiative was launched in 2005 in the Indian subcontinent, the number of cases has come down drastically in this region. However, with the regional incidence of VL decreasing, understanding the role of potentially highly infectious subgroups in maintaining refractory Leishmania transmission is becoming increasingly important. Patients with Post-Kala-Azar Dermal Leishmaniasis (PKDL) are recognized as an important reservoir of transmission. We hypothesized that VL patients co-infected with Human Immunodeficiency Virus (HIV) may also be important reservoirs of sustained leishmania transmission. In this study, we found that the presence of VL-HIV and PKDL patients are both associated with a twofold increase incidence of VL at village level, suggesting they are equally important reservoirs for leishmania transmission. Our paper suggests that VL-HIV patients pose a threat equivalent to PKDL patients towards the VL elimination initiative and that therefore they should receive a similar focus.

## INTRODUCTION

Visceral Leishmaniasis (VL) - also called *kala-azar* (KA) - is a potentially fatal parasitic disease, caused by Leishmania *donovani* on the Indian subcontinent. Fifteen years after a Memorandum of Understanding was signed by the governments of India, Nepal and Bangladesh committing to its elimination as a public health problem, the goal is on the verge of being achieved. However, with the regional incidence of VL decreasing, understanding the role of potentially highly infectious subgroups in maintaining refractory *Leishmania* transmission is becoming increasingly important.

Post kala-azar dermal leishmaniasis (PKDL) is an infectious cutaneous sequel that follows VL in an estimated 5-10% of treated cases in Asia, typically 1-3 years following completion of therapy [1, 2]. Because patients with PKDL usually do not have any symptoms other than painless skin lesions, a minority of these patients actively seek medical care, while the condition is often misdiagnosed as leprosy or vitiligo [1, 3]. Untreated PKDL can remain symptomatic for several years [4, 5]. Following the near elimination of VL from the Indian subcontinent in the 1970s, PKDL was suspected to have been the interepidemic reservoir responsible for triggering a new VL outbreak years after the last VL case had been reported in West Bengal [6]. As such PKDL is considered a largely hidden but persistent reservoir of infection, and remains a major threat to the sustainability of the elimination initiative.

In Bihar, the most endemic state for VL in India, an estimated 2-7% of VL cases are co-infected with HIV [7-9], although this is most likely an underestimation of the true burden [10]. Data from other settings in the Indian subcontinent are limited, in part due to a lack of routine testing [10]. VL patients co-infected with HIV have been shown to be highly infectious [11]. HIV and leishmaniasis share an immunopathological pathway that enhances replication of both pathogens and accelerates the progression of both VL and HIV [12-14]. A concomitant HIV infection increases the risk of developing active VL by between 100 and 2,320 times [15, 16]. Diagnosing VL in HIV-co-infected patients also poses a major challenge, as VL symptoms are less typical and existing diagnostic tools less accurate [17]. In addition, patients with VL-HIV coinfection experience a lower therapeutic success rate for VL, experience greater drug related toxicity and relapse more frequently than patients not infected with HIV [12, 18, 19]. With each new episode of VL becoming increasingly difficult to treat [20], these patients are likely to remain infectious reservoirs for prolonged periods of time. However, their exact contribution to transmission of VL has yet to be determined.

We hypothesized that with the overall decrease in VL cases reported in the Indian subcontinent, PKDL and VL-HIV cases act as important reservoirs of *leishmania* transmission. We therefore investigated to what extent the presence of patients with PKDL or VL-HIV is associated with VL incidence at the village level in Bihar, India.

## METHODS

### Study site

The state of Bihar is located in northern India and is characterized by a very dense population of approximately 120 million, of which over a third live below the poverty line [21]. In 2019, Bihar reported 77% of all VL cases in India [22]. At the same time it carries the second highest number of new HIV infections in the country, and is one of the seven Indian states where reported AIDS related deaths continue to rise [23]. Estimates of the proportion of reported VL patients co-infected with HIV in Bihar state are between 2-7%, although in districts where reliable HIV screening was present up to 20% of adults diagnosed with VL were HIV positive [9, 10]. Six districts within Bihar state were selected for this study on the basis of having the highest VL burden and being geographically contiguous; Darbhanga, Muzaffarpur, Samastipur, Saran, Siwan, and Vaishali (Figure 1).

**Figure 1:**
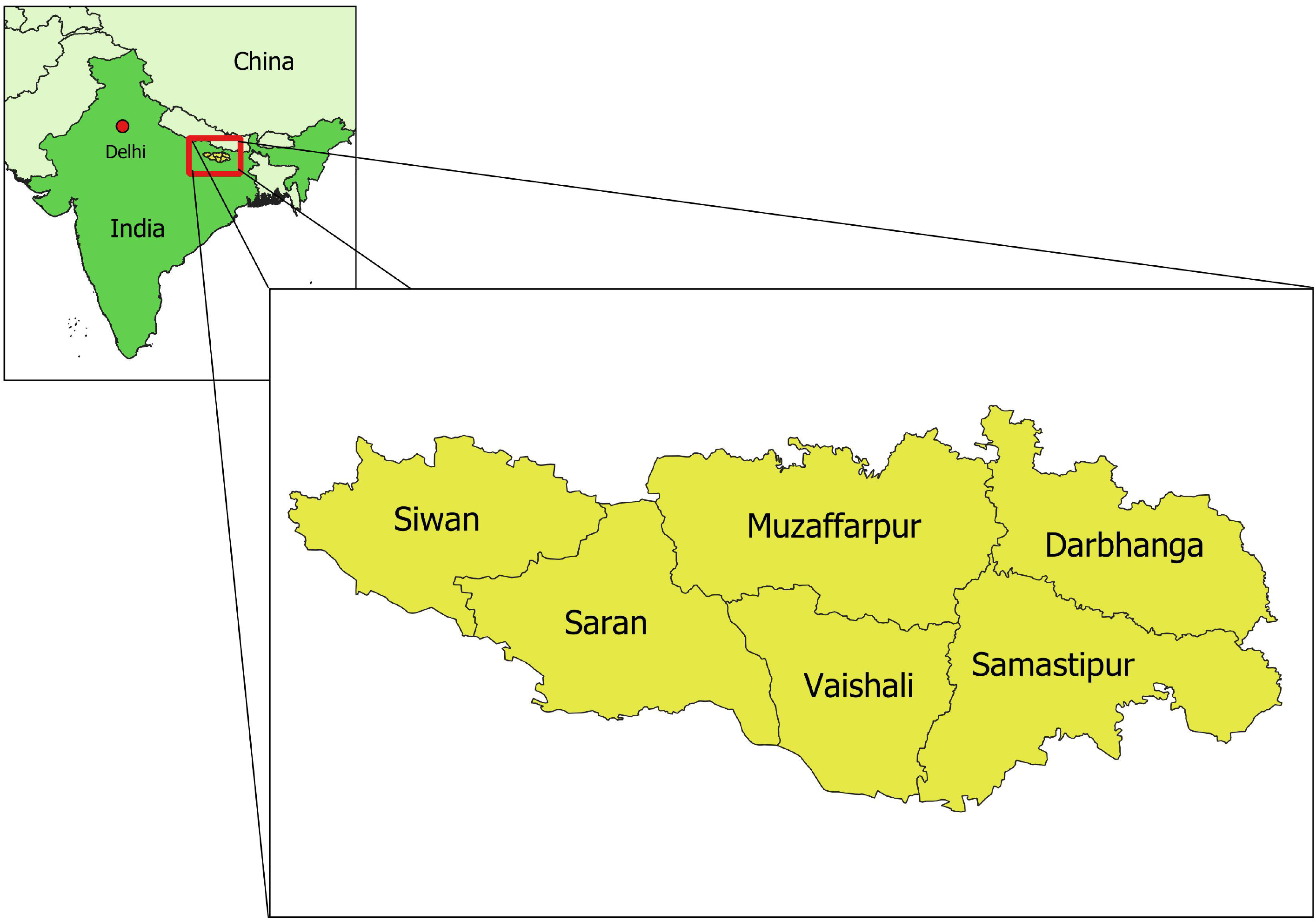
Geographical location of the six study districts within Bihar state, India.

### Data sources

Routinely collected program data – including the date of diagnosis and the village of residence - on incident VL, PKDL and VL-HIV coinfected cases between January 2014 and December 2019 were collected for the six selected districts. These data were retrieved through the Kala-azar Management Information System (KAMIS) – an online reporting tool centralizing case reports of leishmaniasis from all VL endemic states within India - with permission of the National Vector Borne Disease Control Program (NVBDCP), India. The national 2011 population census was used as a basis for the population size per village, and adapted to a 2.5% population growth per year. A village level spatial GIS dataset for these districts was made available through the KalaCORE consortium.

### Data cleaning

The initial database included data on 7,886 patients. Double entries – identified based on date of diagnosis, sex, age, and place of residence - were removed from the database, as were patients for whom the village of residence could not be matched with any of the villages available through the census dataset. VL patients for whom no HIV test result was recorded (n = 2,287) were assumed to be HIV negative. 2,357 villages with less than 1,000 inhabitants were attributed to the nearest larger village, using the ‘distance matrix’ module in Quantum Geographical Information Systems (QGIS). One block (= subdistrict) with no reported cases of VL during the study period was considered to be non-endemic for leishmaniasis and was therefore excluded from the analysis. None of the villages (agglomerates) with a population > 50,000 (n = 9) reported any VL cases; they were therefore excluded from further analysis.

### Statistical analysis

Data analysis was performed with Stata 14 [StataCorp, Texas, USA]. In a first step we assessed whether VL incidence in a village differed by the presence or absence of VL-HIV or PKDL cases. For this we calculated the mean VL incidence in villages with and without:

a. HIV-VL cases in the same year,
b. PKDL cases in the same year,
c. HIV-VL cases in the previous year, and
d. PKDL cases in the previous year, and compared both values using Kruskal-Wallis test (non-normal distribution).

In a next step, we assessed to what extent presence of VL-HIV or PKDL cases was associated with VL incidence at village level, by building a multivariate model using negative binomial regression. The VL-HIV cases themselves were excluded from the outcome variable (VL incidence). As PKDL can be both cause and effect of VL, we evaluated PKDL in the previous year as a potential predictor for VL incidence (instead of PKDL in the same year). Year of reporting and block (= subdistrict) were assessed as potential confounders, block as random effect and year as fixed effect.

### Ethical considerations

Ethical clearance for this study was obtained from the Institutional Review Board of the Institute of Tropical Medicine, Antwerp, Belgium. The retrospective analysis of routinely collected programme data was supported and approved by the National Vector Borne Disease Control Programme (NVBDCP), Ministry of Health and Family Welfare, India.

## RESULTS

### Descriptive statistics

A total of 7,497 cases were included in the analysis; 6,515 cases of VL, 397 cases of VL-HIV, and 585 cases of PKDL. 5.7% (397/6,909) of all VL cases were co-infected with HIV at the time of diagnosis. 37.8% (2,247/5,938) of the included villages reported ≥ 1 case of VL during the study period; while 5.4% (322/5,938) and 7.3% (434/5,938) of the villages reported ≥ 1 HIV-VL case and ≥ 1 PKDL case respectively. Exact distribution of VL, VL-HIV and PKDL cases per year and per district is graphically represented in Figure 2; underlying numbers are available in S1 Table and S2 Table.

**Figure 2:**
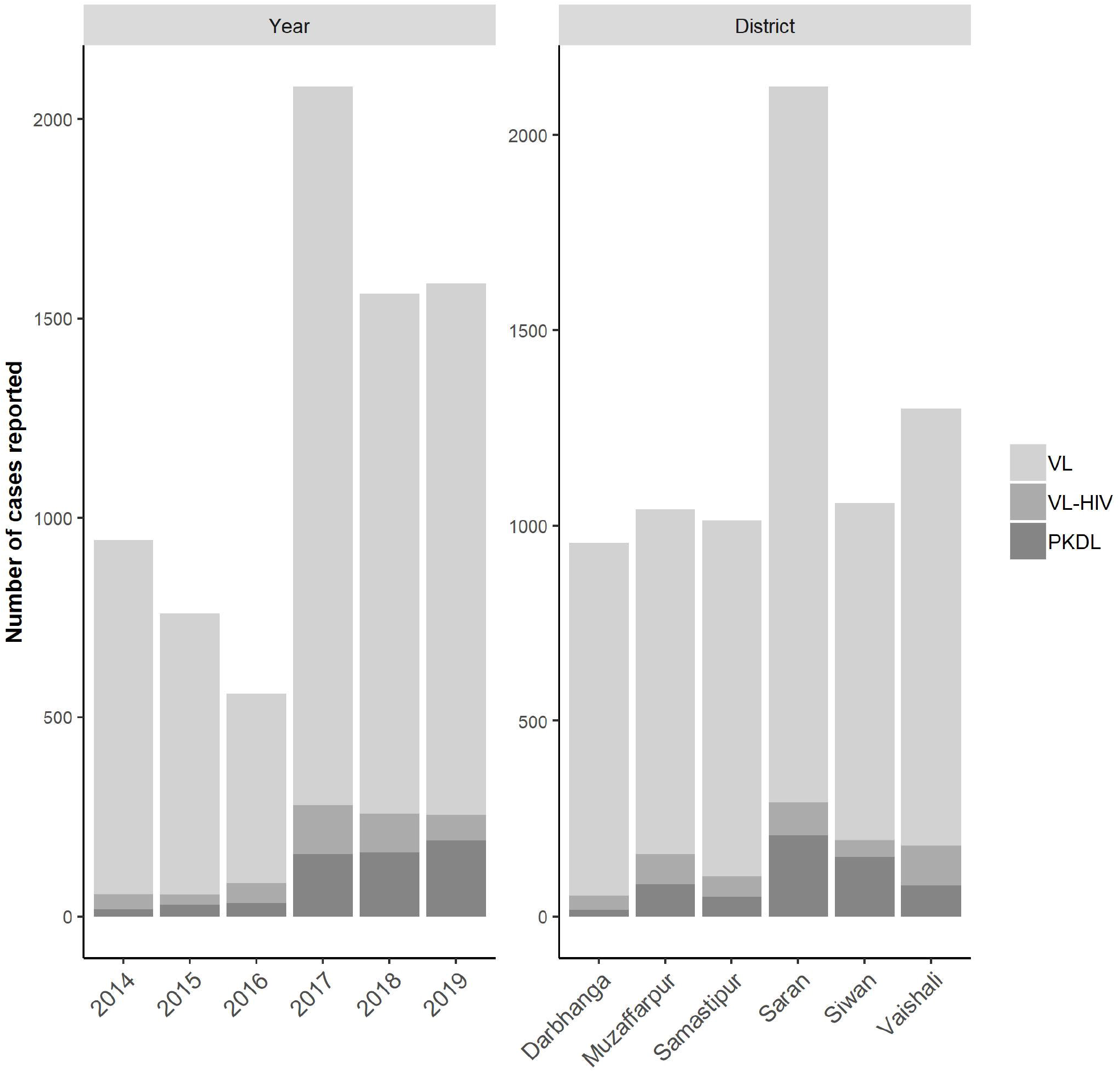
Number of VL, VL-HIV, and PKDL cases reported per year and per district.

### Comparison of VL incidence at village level

The overall mean annual VL incidence of all villages between 2014-2019 was 0.59/10,000 population per year. The mean annual VL incidence rate was significantly higher in villages with VL-HIV or PKDL cases present in the same or the previous year (Table 1). Cumulative incidence per year and per district can be found in S3 Table and S4 Table.

**Table 1:** Mean VL incidence per 10,000 population per year, comparing villages with and without presence of VL-HIV or PKDL cases in the same or the previous year. The reported p-value was calculated using Kruskal Wallis chi square test (non-normal distribution).

|  | Mean VL incidence/10,000/year on village level |  |  |
| --- | --- | --- | --- |
| | present (SD) | absent (SD) | p-value $\chi^2$ |
| **HIV-VL in the same year** | 2.32 (5.70) | 0.58 (2.77) | 0.0001 |
| **PKDL in the same year** | 4.98 (9.98) | 0.53 (2.54) | 0.0001 |
| **HIV-VL in the previous year** | 1.70 (4.70) | 0.68 (3.10) | 0.0001 |
| **PKDL in the previous year** | 2.67 (5.68) | 0.67 (3.07) | 0.0001 |

### Developing the negative binomial model

In the bivariate (unadjusted) analysis, both presence of VL-HIV in the same year and presence of PKDL in the previous year were found to be strong predictors for VL incidence at the village level, with Incidence Rate Ratios (IRR) of 4.07 (95% CI 2.94 – 5.63) and 4.63 (95% CI 3.30 – 6.49) respectively. The year of reporting was identified as a confounder.

A multivariate model was then built, with VL incidence as outcome, and the presence of VL-HIV cases in the same year as well as presence of PKDL cases in the previous year as predictors, while controlling for year as a confounder and using block as a random effect to adjust for contextual confounding. The model was further explored by adding an interaction term for VL-HIV(same year)*PKDL(previous year), but no interaction was found to be present. Our final model can therefore be summarized as follows: VL incidence ∼ VL-HIV (same year) + PKDL (previous year) + year + block(random effect). In summary, presence of VL-HIV in the same year and of PKDL in the previous year both remained strong predictors for VL incidence at village level, with an adjusted IRR of 2.16 (95% CI 1.81 – 2.58) and 2.37 (95% CI 2.01 – 2.81) respectively. Results of the bivariate (unadjusted) and multivariate (adjusted) negative binomial regression model can be found in Table 2.

**Table 2:**
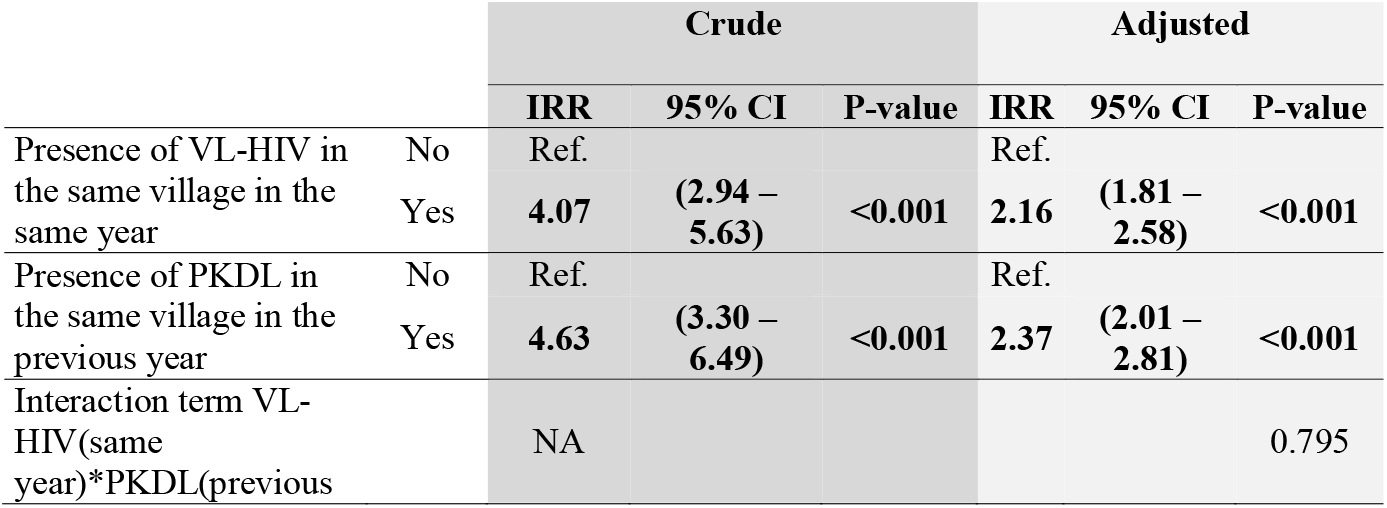
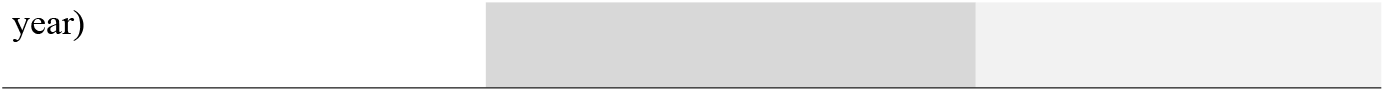
Results of bivariate (crude) and multivariate (adjusted) analysis using negative binomial regression. Incidence rate ratios (IRR) for VL are presented, including 95% confidence intervals (95% CI) and p-values. Statistically significant associations (p-values <0.05) are highlighted in bold.

### Sensitivity analysis

A sensitivity analysis was done fitting the final multivariate negative binomial model for each of the districts separately. As shown in Figure 3, this consistently showed an increased VL incidence in the presence of current VL-HIV cases or PKDL cases in the previous year in all districts, though for PKDL this difference was not significant in one district (underlying numbers and confidence intervals are detailed in S5 Table).

**Figure 3:**
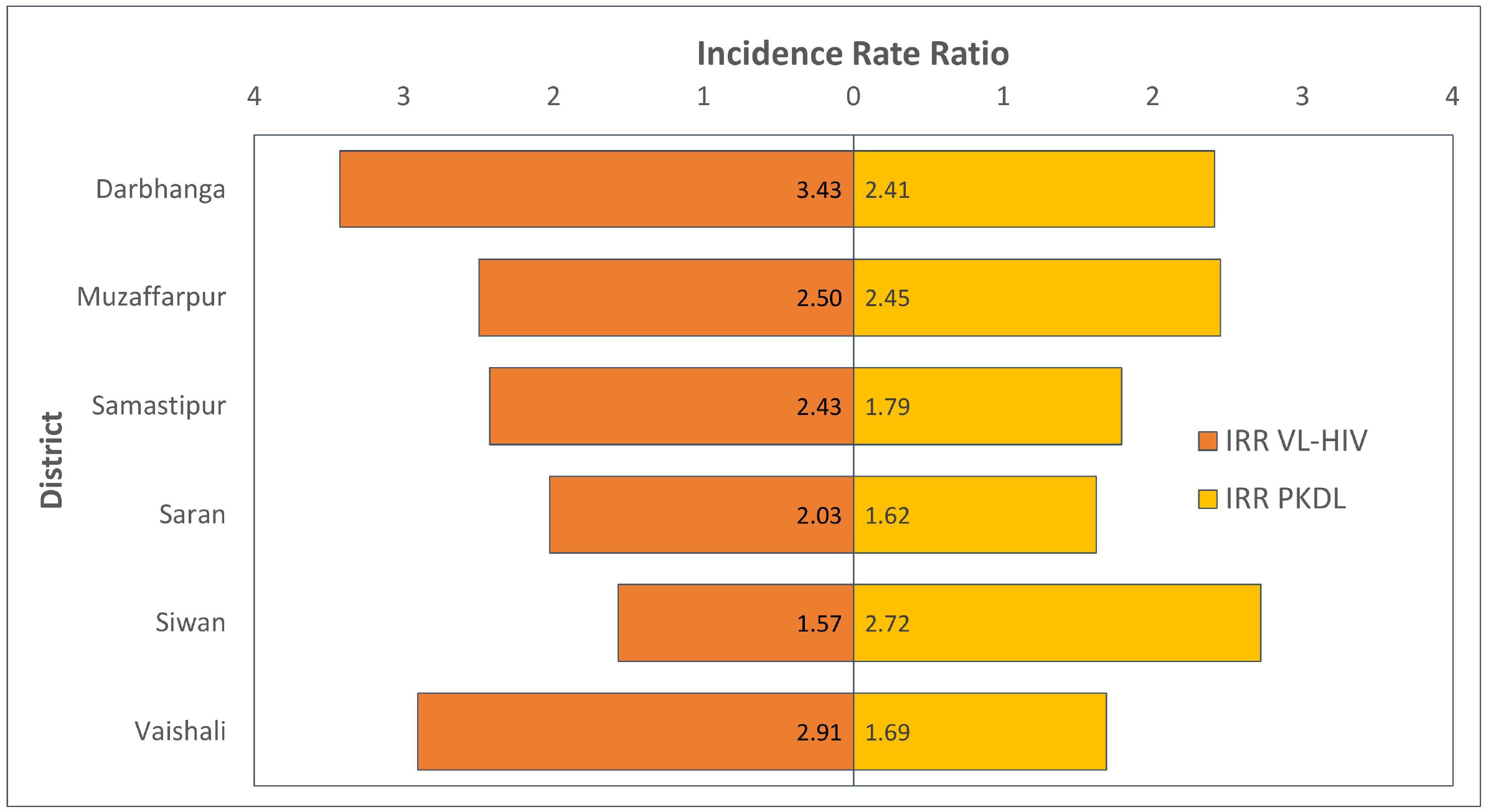
Tornado chart graphically representing the results of the sensitivity analysis of the final model for the different districts. On the horizontal axis, the adjusted Incidence Rate Ratio is displayed for VL-HIV in the same year and PKDL in the previous year respectively.

## DISCUSSION

This study suggests that presence of either VL-HIV or PKDL patients is associated with a greater than twofold increase in VL incidence at village level (IRR 2.16 and 2.37 for VL-HIV and PKDL respectively). A similar effect of VL-HIV and PKDL was found in all six districts; though not statistically significant in all, demonstrating a consistent trend supports causality as an explanation. Although the case for focus on PKDL identification and treatment is widely accepted as an important strategy in achieving sustainable elimination [1], these findings indicate the importance of VL-HIV patients as infectious reservoirs, and that a lack of equivalent focus on this cohort may represent a similar level of threat to the VL elimination initiative on the Indian subcontinent.

This is the first study that attempts to quantify the impact of VL-HIV on population-level transmission within endemic areas. The main strength of this analysis is that our model takes into account the chronological hierarchy between VL and PKDL, excluding as a predictor the PKDL cases which might have developed in a VL case reported earlier in the same year, which would artificially have increased the association.

There are a number of limitations in this study. Routinely collected programme data will likely be an underestimation of the real disease burden, not only of VL but also of VL-HIV co-infections and PKDL, the latter two of which are difficult to identify. Despite active case finding and numerous information, education and communication (IEC) initiatives implemented as part of the elimination strategy, PKDL patients tend not to seek medical care unless for cosmetic reasons, as they do not feel unwell. Simultaneously, the recommendation for routine testing for HIV of all patients diagnosed with VL in India was only made in 2014, and has taken time to be implemented. Meanwhile, there is no routine screening for VL in patients with HIV living in VL endemic areas in India, despite national recommendations, primarily because of the lack of tools to enable this [9].

Additionally, in the analysis, VL patients for whom no HIV test result was recorded were assumed to be HIV negative. Zero reporting of results in health care surveillance systems remains uncommon, so although this assumption will most likely be correct for most of the patients, some VL cases may have been incorrectly considered as HIV negative.

Both PKDL and VL-HIV patients have been shown to be infectious through xenodiagnosis, a procedure in which a vector is allowed to feed on a host in order to verify whether or not they get infected. A recent study from Bangladesh showed that PKDL is only slightly less infectious than VL, with 57% of PKDL patients infecting at least one sand fly compared to 67% of VL patients [24]. Through the same technique, patients with VL-HIV have also been shown to be highly infectious, with even asymptomatic cases infecting vectors, albeit with a different *leishmania* species and in a different context [25, 26]. However no such study on VL-HIV patients has been published on the Indian subcontinent.

Evidence on the relative contributions of PKDL and VL-HIV to the transmission of *leishmania* is still evolving. While a 2016 longitudinal study from India reported no evidence of increased transmission in households with PKDL patients over an 18-month follow-up period compared to control households [27], mathematical modeling suggests an increasingly important role for PKDL in the years to come [28]. VL-HIV cases on the other hand are traditionally not included in mathematical models, and the results of this study suggest that this be an important area of consideration for modelers. While the necessity to include PKDL in the elimination efforts has by now been widely accepted, this study suggests that the VL-HIV reservoir warrants an equivalent focus in operational strategies if sustainable elimination is to be achieved. The results of this study would *a-priori* also support the recent recommendation of the use of LLINs in patients diagnosed with PKDL and VL-HIV from the 7^th^ WHO-SEARO Regional Technical Advisory Group for Kala Azar.

There remain major gaps in the diagnosis and management of both PKDL and VL-HIV in the Indian subcontinent. PKDL remains for the most part a clinical diagnosis, with the potential for mis-identification and missed treatment remaining a major issue. Meanwhile, treatment of PKDL remains challenging, with a 12-week course of oral Miltefosine, a teratogenic oral drug with poor compliance and significant side effects, being the only currently recommended treatment [29]. Meanwhile, there remains little evidence on the sensitivity or specificity of existing non-invasive tests for VL in HIV patients, and considering the case definition of VL remains the same for co-infected and immunocompetent patients, it is likely that a substantial number of co-infected patients are missed. In addition, management of co-infected patients remains fraught with challenges [10, 30]. New tools and strategies are urgently required to improve case detection and management of both entities to reduce their infectious potential. This will both improve the chances, and indeed capitalize on the substantial investment, of sustainable elimination in the subcontinent.

## Data Availability

Data cannot be shared publicly by the researchers as they are property of the National Vector Borne Disease Control Programme (NVBDCP) of India. They are available to others on condition of request and approval of the NVBDCP (contact via).

## Author contributions

EH, SB, PM, and KC contributed to the conception and design of the study. Data curation was performed by NG, PM and EH. Formal analysis was performed by PM and KC and validated by SB and EH. Writing of the original draft manuscript was performed by PM and KC. All authors contributed to manuscript revision, read, and approved the submitted version.

## REFERENCES

1. Zijlstra EE, Alves F, Rijal S, Arana B, Alvar J. Post-kala-azar dermal leishmaniasis in the Indian subcontinent: A threat to the South-East Asia Region Kala-azar Elimination Programme. PLoS neglected tropical diseases. 2017;11(11):e0005877.

2. Zijlstra EE, Musa AM, Khalil EA, el-Hassan IM, el-Hassan AM. Post-kala-azar dermal leishmaniasis. Lancet Infect Dis. 2003;3(2):87–98.

3. Ramesh V, Kaushal H, Mishra AK, Singh R, Salotra P. Clinico-epidemiological analysis of Post kala-azar dermal leishmaniasis (PKDL) cases in India over last two decades: a hospital based retrospective study. BMC Public Health. 2015;15:1092.

4. Das AK, Harries AD, Hinderaker SG, Zachariah R, Ahmed B, Shah GN, et al. Active and passive case detection strategies for the control of leishmaniasis in Bangladesh. Public Health Action. 2014;4(1):15–21.

5. Garapati P, Pal B, Siddiqui NA, Bimal S, Das P, Murti K, et al. Knowledge, stigma, health seeking behaviour and its determinants among patients with post kalaazar dermal leishmaniasis, Bihar, India. PloS one. 2018;13(9):e0203407.

6. Addy M, Nandy A. Ten years of kala-azar in west Bengal, Part I. Did post-kala-azar dermal leishmaniasis initiate the outbreak in 24-Parganas? Bull World Health Organ. 1992;70(3):341–6.

7. Mathur P, Samantaray JC, Vajpayee M, Samanta P. Visceral leishmaniasis/human immunodeficiency virus co-infection in India: the focus of two epidemics. J Med Microbiol. 2006;55(Pt 7):919–22.

8. Burza S, Mahajan R, Sanz MG, Sunyoto T, Kumar R, Mitra G, et al. HIV and visceral leishmaniasis coinfection in Bihar, India: an underrecognized and underdiagnosed threat against elimination. Clinical infectious diseases : an official publication of the Infectious Diseases Society of America. 2014;59(4):552–5.

9. Directorate National Vector Borne Disease Control Programme India. Accelerated plan for Kala-azar elimination 2017 [Available from: https://nvbdcp.gov.in/WriteReadData/l892s/Accelerated-Plan-Kala-azar1-Feb2017.pdf] (accessed May 10, 2020).

10. Akuffo H, Costa C, van Griensven J, Burza S, Moreno J, Herrero M. New insights into leishmaniasis in the immunosuppressed. PLoS neglected tropical diseases. 2018;12(5):e0006375.

11. Molina R, Gradoni L, Alvar J. HIV and the transmission of Leishmania. Annals of tropical medicine and parasitology. 2003;97 Suppl 1:29–45.

12. Alvar J, Aparicio P, Aseffa A, Den Boer M, Canavate C, Dedet JP, et al. The relationship between leishmaniasis and AIDS: the second 10 years. Clinical microbiology reviews. 2008;21(2):334–59.

13. Tremblay M, Olivier M, Bernier R. Leishmania and the pathogenesis of HIV infection. Parasitology today (Personal ed). 1996;12(7):257–61.

14. Mock DJ, Hollenbaugh JA, Daddacha W, Overstreet MG, Lazarski CA, Fowell DJ, et al. Leishmania induces survival, proliferation and elevated cellular dNTP levels in human monocytes promoting acceleration of HIV co-infection. PLoS pathogens. 2012;8(4):e1002635.

15. Lopez-Velez RP-MJAGABFVJELBCP-CFAJ. Clinicoepidemiologic characteristics, prognostic factors, and survival analysis of patients coinfected with human immunodeficiency virus and leishmania in an area of Madrid, Spain. 1998.

16. World Health Organization. Leishmaniasis and HIV coinfection [Available from: https://www.who.int/leishmaniasis/burden/hiv_coinfection/burden_hiv_coinfection/en/] (accessed April 7, 2020).

17. Singh S. Changing trends in the epidemiology, clinical presentation, and diagnosis of Leishmania-HIV co-infection in India. International journal of infectious diseases : IJID : official publication of the International Society for Infectious Diseases. 2014;29:103–12.

18. Cota GF, de Sousa MR, Rabello A. Predictors of Visceral Leishmaniasis Relapse in HIV-Infected Patients: A Systematic Review. PLoS neglected tropical diseases. 2011;5(6):e1153.

19. Burza S, Mahajan R, Sinha PK, van Griensven J, Pandey K, Lima MA, et al. Visceral Leishmaniasis and HIV Co-infection in Bihar, India: Long-term Effectiveness and Treatment Outcomes with Liposomal Amphotericin B (AmBisome). PLoS neglected tropical diseases. 2014;8(8):e3053.

20. van Griensven J, Carrillo E, Lopez-Velez R, Lynen L, Moreno J. Leishmaniasis in immunosuppressed individuals. Clinical microbiology and infection : the official publication of the European Society of Clinical Microbiology and Infectious Diseases. 2014;20(4):286–99.

21. The World Bank. Bihar Poverty, Growth and Inequality. 2016 [Available from: http://documents.worldbank.org/curated/en/781181467989480762/pdf/105842-BRI-P157572-PUBLIC-Bihar-Proverty.pdf] (accessed May 20, 2020).

22. National Vector Borne Disease Control Programme I. National Vector Borne Disease Control Programme, India. Kala-azar Cases and Deaths in the country since 2013. [Available from: https://nvbdcp.gov.in/index4.php?lang=1&level=0&linkid=467&lid=3750] (accessed March 23, 2020).

23. National AIDS Control Organization, India. India HIV estimations 2017 - fact sheets [Available from: http://naco.gov.in/sites/default/files/HIV%20Estimations%202017%20Report_1.pdf] (accessed April 5, 2020).

24. Mondal D, Bern C, Ghosh D, Rashid M, Molina R, Chowdhury R, et al. Quantifying the Infectiousness of Post-Kala-Azar Dermal Leishmaniasis Toward Sand Flies. Clinical infectious diseases : an official publication of the Infectious Diseases Society of America. 2019;69(2):251–8.

25. Molina R, Lohse JM, Pulido F, Laguna F, López-Vélez R, Alvar J. Infection of sand flies by humans coinfected with Leishmania infantum and human immunodeficiency virus. The American journal of tropical medicine and hygiene. 1999;60(1):51–3.

26. Ferreira GR, Castelo Branco Ribeiro JC, Meneses Filho A, de Jesus Cardoso Farias Pereira T, Parente DM, Pereira HF, et al. Human Competence to Transmit Leishmania infantum to Lutzomyia longipalpis and the Influence of Human Immunodeficiency Virus Infection. The American journal of tropical medicine and hygiene. 2018;98(1):126–33.

27. Das VN, Pandey RN, Siddiqui NA, Chapman LA, Kumar V, Pandey K, et al. Longitudinal Study of Transmission in Households with Visceral Leishmaniasis, Asymptomatic Infections and PKDL in Highly Endemic Villages in Bihar, India. PLoS neglected tropical diseases. 2016;10(12):e0005196.

28. Le Rutte EA, Zijlstra EE, de Vlas SJ. Post-Kala-Azar Dermal Leishmaniasis as a Reservoir for Visceral Leishmaniasis Transmission. Trends in parasitology. 2019;35(8):590–2.

29. Pijpers J, den Boer ML, Essink DR, Ritmeijer K. The safety and efficacy of miltefosine in the long-term treatment of post-kala-azar dermal leishmaniasis in South Asia - A review and meta-analysis. PLoS neglected tropical diseases. 2019;13(2):e0007173.

30. Cota GF, de Sousa MR, Fereguetti TO, Rabello A. Efficacy of anti-leishmania therapy in visceral leishmaniasis among HIV infected patients: a systematic review with indirect comparison. PLoS neglected tropical diseases. 2013;7(5):e2195.

